# Patients’ perceptions of, emotional responses to, and disengagement from, physician-services for HIV: The relationship of trust and the mental and social effects of HIV to perceived service-adequacy

**DOI:** 10.1101/2024.05.16.24307496

**Authors:** R Whitaker, M Otis

## Abstract

An exploratory, two-language, online survey assessed patients in relation to physician-services for HIV-infection. Most respondents had experienced primarily mental rather than physical illness due to HIV or its treatment, as well as a marked HIV-related impact on social networks. Mental and social factors were identified that predicted patients’ relationship to, and disengagement from, physicians’ services, including patients’ satisfaction, trust, physicians’ relational skills, time-pressure, the opportunity for clinical intimacy, physician-credibility; ‘heart-sink’ physicians, anxiety, passivity, self-efficacy, experience of HIV-related illness, experience of medication-toxicity, degree of loss in social networks, and ‘changing physician due to disagreement’, with ramifications for physicians’ concept of the ‘difficult patient’. The prevalence and significance of such issues and the related risk of distrust of, and disengagement from, physician-services confirm the still-remaining need for evidence-based treatment to be implemented for people with HIV, to include the delivery of integrated, multi-professional, and biopsychosocial health-services.

**What do we know already about this topic?:** Mental and social aspects of illness affect services’ ability to help patients to get well and stay well (i.e., health-effectiveness), but since the advent of HAART few clinics provide integrated, biopsychosocial services necessary to address these issues, despite the extensive evidence-base demonstrating such need.

**How does this research contribute to the field?:** This research: confirms the significance of biopsychosocial aspects of illness and identifies pervasive factors that intensify such challenges to the provision of health-effective services—and which can act as signal-events in quality-related service-audit—as well as analysing other factors, such as patients’ satisfaction, that have no value except to consumerism but relate to other variables, such as trust, that need to be used in service-evaluation.

**What are the implications of this research for theory, practice, or policy?:** The implications of this research are that each services needs a competent, multi-professional team to address the biopsychosocial factors identified, and to integrate the factors into the audit of services’ health-effectiveness, supported by specific policy such as treatment-guidelines that require the evidence-based formulation and delivery of health-services, not solely pharmaceuticals of choice.

## Introduction

The primary goal of all health-services should be to maximise patients’ health, as defined by the World Health Organization^1^. In order to achieve sustainable increases in health, patients’ commitment to treatment and their health-goals is critical^2^. Understanding patients’ relationships to medical services is key to maximising such commitment and to such services’ ability to be health-effective. It is important to find scientifically valid indices of patients’ relationships to medical services that have a strong predictive relationship to comprehensive, sustainable health-outcomes, thus demonstrating services’ health-effectiveness. Patient-satisfaction as a construct has emerged from American approaches to patients as consumers^3^ and is currently used as a quality-index; however, there is no scientific evidence demonstrating any adequate predictive value of satisfaction in relation to a service’s quality and health-effectiveness.

Patients’ commitment to treatment requires a ‘buy-in’^4^ that occurs following a conditional permission to receive medical services, an acknowledgement of the utility and value of the service, consent to disease- and illness-treatments, an endorsement of a treatment-plan, and a motivated engagement in that treatment through whatever difficulties might arise. Failures in services’ achievement or sustaining of patient-commitment are reflected in poor uptake and retention of services by patients — called “linkage to care” from a physician-centred perspective — which are of particular concern in HIV-disease^5^. Failures to address the determinants of commitment and trust^6^ are similarly reflected in complaints, malpractice-litigation, and other problems that have significance for performance-based financing of services. In HIV-infection, given the importance of lifelong commitment to medical treatment by patients, we explore some of the aspects of patients’ relationships to physician-services for HIV-infection that are plausible determinants of sustained patient-commitment to treatment, as well as their relationship to patient-satisfaction.

## Methods and Sample-Characteristics

### Development and delivery

An exploratory online survey was used to examine people’s relationships to physician-services for HIV-infection. The survey was designed by the first author, a clinician-scientist with long-term HIV-infection, through literature-research and patient-interviews using approaches from grounded theory. The self-inclusion criteria for this study were being HIV-positive, being of ten years of age or older, and having access to physician-services for HIV-disease. The Human and Life Sciences sub-committee review-board at Roehampton University, UK, provided ethical approval of the survey’s methodology. The survey was hosted and promoted through www.thebody.com, an international, online portal translating biomedical perspectives on HIV-disease to people with HIV-infection. The survey was available in English and Spanish, with the English original being translated into Spanish by native Spanish-speakers and back-translated into English by native English-speakers to ensure validity.

The survey consisted of 41 questions with approximately 450 nested response-options addressing sociodemographic, attitudinal, and clinical variables, including accessibility and utilisation of health-services. Only relevant, contingent response-options were revealed to respondents, minimising the perceived scale of the survey; optional feedback reported uniformly that the questionnaire was simple and easy to complete.

### Geographic and sociodemographic characteristics of the sample

Geographic and sociodemographic questions included age, sexual orientation, current place of residence, where the respondent grew up (from birth to 20 years of age), ethnic-group identification (specified by geographic area first and then the names of ethnic groups as used in that area—e.g., ‘African-American’ in the USA versus ‘Black African’ or ‘Black Caribbean’ in the UK), and language used to answer the survey. We estimated migration-status by comparing where the person grew up to where they live now. Data as to residence, ethnicity, migration, age, gender, and sexual orientation are presented in Tables 1-2.

**Table 1:** Residence, ethnicity, and migration-characteristics of the sample.

| Characteristic | N | %age |
| --- | --- | --- |
| **Current residence** | n=368 | 87.0 (of total) |
| **The Americas** | **247** | **58.4** |
| North | 172 | 46.7 |
| South | 26 | 7.1 |
| Central/ Mexico | 19 | 5.2 |
| **Europe** | **88** | **23.9** |
| West | 45 | 12.2 |
| South/ South-East | 15 | 4.1 |
| Central | 12 | 3.3 |
| North | 6 | 1.6 |
| **Asia or the Caucasus** | **13** | **3.5** |
| South-East | 8 | 2.2 |
| **Australasia / Oceania** | **7** | **1.9** |
| Australia | 6 | 1.6 |
| **Africa** | **3** | **0.8** |
| **Middle East** | **2** | **0.5** |
| **Caribbean** | **2** | **0.5** |
| **Other\*** | **6** | **1.6** |
| **Ethnicity by residence** | n=218 | 51.5 (of total) |
| **North American** | **85** | **39.0** |
| Asia/Caucasian | 39 | 17.9 |
| European | 25 | 11.5 |
| Hispanic | 9 | 4.1 |
| Other | 12 | 5.5 |
| **European** | **70** | **32.1** |
| British/ Irish | 27 | 12.4 |
| South Eastern | 22 | 10.1 |
| Central/ Northern | 15 | 6.9 |
| Other | 6 | 2.8 |
| **South/ Central American** | **63** | **28.9** |
| Hispanic | 23 | 14.7 |
| Other | 5 | 2.3 |
| **Other** | **35** | **16.1** |
| **Migration: direction** | n=75 | 17.7 (of total) |
| Americas to Europe | 5 | 6.7 |
| Middle East to Europe | 2 | 2.6 |
| Europe to Americas | 4 | 5.3 |
| Caribbean to Americas | 1 | 1.3 |
| Asia to Americas | 1 | 1.3 |
| Other to Americas | 3 | 4.0 |
| Europe to Asia | 1 | 1.3 |
| Europe to Australasia | 1 | 1.3 |
| No direction identifiable | 57 | 76.0 |

**Table 2:** Sociodemographic characteristics of the sample.

| Characteristic | N | %age |
| --- | --- | --- |
| **Age-range (years)** | n=423 |  |
| 10-19 | 6 | 1 |
| 20-29 | 49 | 12 |
| 30-39 | 116 | 28 |
| 40-49 | 155 | 37 |
| 50-59 | 71 | 17 |
| 60+ | 26 | 6 |
| **Current Gender \| Sexual Orientation** | n=423 |  |
| **Male (sub-total n)** | **377** | **89.1** |
| Gay | 325 | 76.8 |
| Heterosexual | 33 | 7.8 |
| Bisexual | 19 | 4.5 |
| **Female (sub-total n)** | **46** | **10.9** |
| Heterosexual | 40 | 9.5 |
| Bisexual | 2 | 0.5 |
| Lesbian | 4 | 1.0 |

The sample consisted of 423 respondents with 23 regionally specified ethnicities from more than 16 countries and all inhabited continents; overall, the survey-results had a 4.9% margin of error. The sample showed a broad range of ages and all acknowledged sexual orientations, though fewer women compared to men, fewer heterosexual men compared to gay men, and negligible numbers of people younger than 20 and over 59, lesbians, and bisexual men and women. While current gender was asked, neither sex at birth or transgender status was assessed, which was a signal failing in design. Further, due to a human-factors’ error with the entry-screen to the survey, language-data were not properly available; however, of the 44% (n=188) respondents in which it was possible to identify the language used to answer the survey through free-text responses, 79% (n=149) used the English-language interface and 21% (n=40) the Spanish-language interface. Of the latter, 73% (n=29) were from South America, 23% (n=9) from Europe, and only 2.4% (n=1) from North America, a distribution that was unexpected.

Overall, most respondents were from the USA and English-speaking. Of those whose migration-status could be estimated from the country in which they were born compared to the country where they were currently resident, only 5% were migrants and no particular geographical direction of migration was notable.

### Clinical characteristics of the sample

Data as to clinical characteristics of the sample are shown in Table 3. There was a good representative range of participants dependent on how long they had known they have HIV, ranging from ‘less than one year’ to ‘over 20 years’. Only one person reported having been infected from birth; those aged 10-19 were most likely to have been infected for 0-2 years. Length of infection increased with age-group, except for those over 70 years of age (n=4) who reported having been infected for 3-5 years or 11-20 years. The sample showed a monotone decrease in terms of length of time with HIV-infection. Furthermore, 71.4% (302/423) reported ever having taken HIV-medication (antiretrovirals); we also asked specifically about medication-toxicity (i.e., negative side-effects), adherence; the analysis of adherence-related factors will be reported separately. Finally, we asked how many times the respondent had changed physicians due to a disagreement, which showed that almost a third of the sample had done so.

**Table 3:** Clinical characteristics of the sample.

| Characteristic | N | %age |
| --- | --- | --- |
| Length of time with HIV-infection | n=423 |  |
| Less than a year | 69 | 16.3 |
| 1-2 years | 82 | 19.4 |
| 3-5 years | 99 | 23.4 |
| 6-10 years | 69 | 16.3 |
| 11-20 years | 65 | 15.4 |
| More than 20 years | 39 | 9.2 |
| Time taking medication for HIV-infection | n=423 |  |
| Never | 121 | 28.6 |
| Less than a year | 68 | 16.1 |
| 1-2 years | 64 | 15.1 |
| 3-5 years | 76 | 18.0 |
| 6-10 years | 42 | 9.9 |
| 11-20 years | 52 | 12.3 |
| Times changed medication \| toxicity | n=302 | 71.4 |
| Never | 164 | 54.3 |
| 1 | 70 | 23.2 |
| 2 | 37 | 12.2 |
| 3 | 16 | 5.3 |
| 4 | 6 | 2.0 |
| 5 | 3 | 1.0 |
| >5 | 6 | 2.0 |
| Times changed physician \| disagreement | n=357 | 84.4 |
| Never | 247 | 69.2 |
| 1 | 74 | 20.7 |
| 2 | 26 | 7.3 |
| 3 | 6 | 1.7 |
| 4 | 4 | 1.1 |

As the sample’s characteristics overall were diverse, we determined its possible generalisability by computing rank-order correlations between percentages in strata defined by age (under 40 versus over 40), region (North America, South America, Britain, Europe, and Other), and years since learning of their HIV infection (within the last year, 1-2 years ago, 3-5 years ago, and 6 or more years ago), as shown in Table 4. The correlations suggest that samples of different compositions of age, region, and number of years of known HIV-infection would yield similar relative magnitudes of prevalence across socio-demographic options, albeit with some exceptions (e.g., low-value coefficients between “Other” region and Britain and between recent knowledge of HIV infection versus 1-2 years), indicating that this sample is not inconsistent statistically with a proper random sample.

**Table 4:** Pearson-correlations for each cross-strata, rank-order comparison.

| Strata | Average r | Range of r |
| --- | --- | --- |
| 2 strata: age (1 correlation) | .931 | --- |
| 5 strata: region (10 correlations) | .647 | .240 to .896 |
| 4 strata: #years known HIV+ (6 correlations) | .703 | .494 to .888 |

Respondents were asked if they had ever had any physical or emotional problems due to HIV or its treatment, with response-options allowing the reporting of mild, moderately serious, serious, and very serious symptoms, and the number of each such experience within each category. Of the total respondents, 267 (63%) reported such problems. Further, 33% (n=137) of all respondents reported having had “physical or emotional problems because of another chronic medical condition *unrelated* to HIV or its treatment”; of these, 17.5% (n=24) reported ‘severe’ or ‘very severe’ problems. Significantly, the problems most reported were mental disorders (61%, n=90), specifically: depression (42%, n=62); anxiety or stress-disorders (8.1%, n=12); behavioural or chemical dependencies (8.1%, n=12); and cognitive disorders (2.7%, n=4).

The next highest were liver-related problems (13.6%, n=20; particularly due to Hepatitis C co-infection, including two liver-transplants); gastrointestinal problems (9.5%, n=14), particularly diarrhoea; musculoskeletal problems (8.8%, n=13); and metabolic problems, including hormonal (thyroid, adrenal, pancreatic), and fat- or muscle-wasting (8.1%, n=12).

However, textual responses showed that respondents did not or could not distinguish between conditions that were more likely to be due to HIV-infection or its treatment (e.g., osteopaenia, neuropathy, lipoatrophy, cytomegalovirus-infection), those that may or may not have been (depression, cardiovascular disease, diabetes), and those that were likely to have been biologically and causatively distinct, although they may have occurred through, or acted as, biological, behavioural, or biobehavioural factors in common with the acquisition or course of HIV-infection (e.g. Hepatitis B or C infections, Attention-Deficit and Hyperactivity Disorder, degenerative disc-disease prior to diagnosis with HIV-infection). This may reflect problems in differential diagnosis, communication by physicians, and/or respondents’ abilities to understand.

For the sake of easier comprehension, we created a composite variable of the personal experience of HIV-illness due to HIV or HIV-treatment, with purposefully subjective but spline ordinal response-options. The response-options were weighted 1-4 in order, and then multiplied by the incidence of problems reported for each weighted item; the resulting distribution of 40 data-points was cut into 6 groups and the distribution determined, to create an HIV-illness score based on severity and incidence (Table 5).

**Table 5:** Experience of HIV-illness Composite of Severity by Incidence.

| Items included | Composition | Descriptives |
| --- | --- | --- |
| 1. Have you ever had any physical or emotional problems due to HIV or its treatment?<br>a. No<br>b. If yes, how many problems have you had that were **mild**?<br>c. If yes, how many problems have you had that were **moderately serious**?<br>d. If yes, how many problems have you had that were **serious**?<br>e. If yes, how many problems have you had that were **very serious**? | 1. Response-options: 1= "one or a few", 2= "more than a few but not many", 3= "Quite a lot", 4= "A huge number"<br>2. The responses as to number of problems were weighted by severity (mild=1; moderately serious=2; serious=3; very serious=4) and summed<br>3. The summed responses were collapsed into 6 groups | Mean=1.5<br>SD=1.3<br>n=422<br>Range=0-5 |
| Scores | n=423 | %age |
| 0 | 156 | 36.9 |
| 1-5 | 91 | 21.8 |
| 6-10 | 58 | 13.7 |
| 11-15 | 64 | 15.1 |
| 16-25 | 33 | 7.8 |
| 26-40 | 20 | 4.7 |

### Impact on social networks

This variable consisted of the respondent’s estimation of the number of people known to have been sick or to have died among those who were “close or family” or “less-close” to the respondent. Figure 1 shows the descriptive statistics of these responses. A majority of the respondents reported knowing others who had been sick or died from HIV (85%, n=305) but a significant minority had not (15%, n=52).

**Figure 1:**
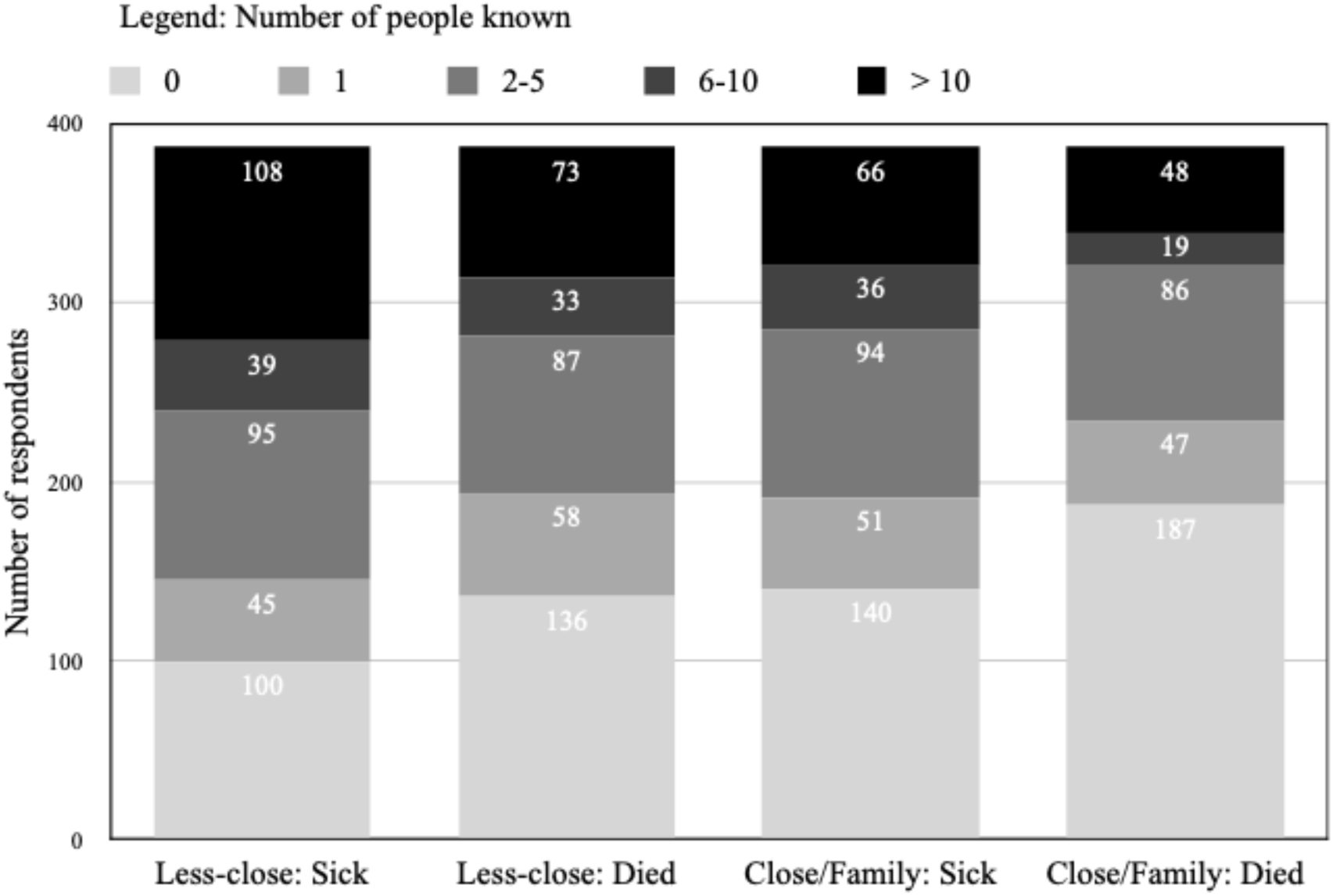
Impact on social network: Proximity-by-severity.

There was significant variation among the sample: 10.1% of the overall sample had known more than 50 people “less close” to them to have been sick and 6.8% had known this many people to have died; 15.6% had known more than 10 family-members or people close to them to have been sick and 11.3% knew more than 10 such people to have died. We also created a composite variable of others’ HIV-illness in their social networks, as an index of the impact on the respondents’ social networks, in which ‘social distance’ was weighted 1-4, from ‘less-close and sick’, ‘less-close and died’, ‘close/family and sick’, ‘close/family and died’, and applied to the incidence in each category, with the resulting distribution of 61 data-points cut into 9 groups and the distribution characterised (Table 6; Figure 2).

**Figure 2:**
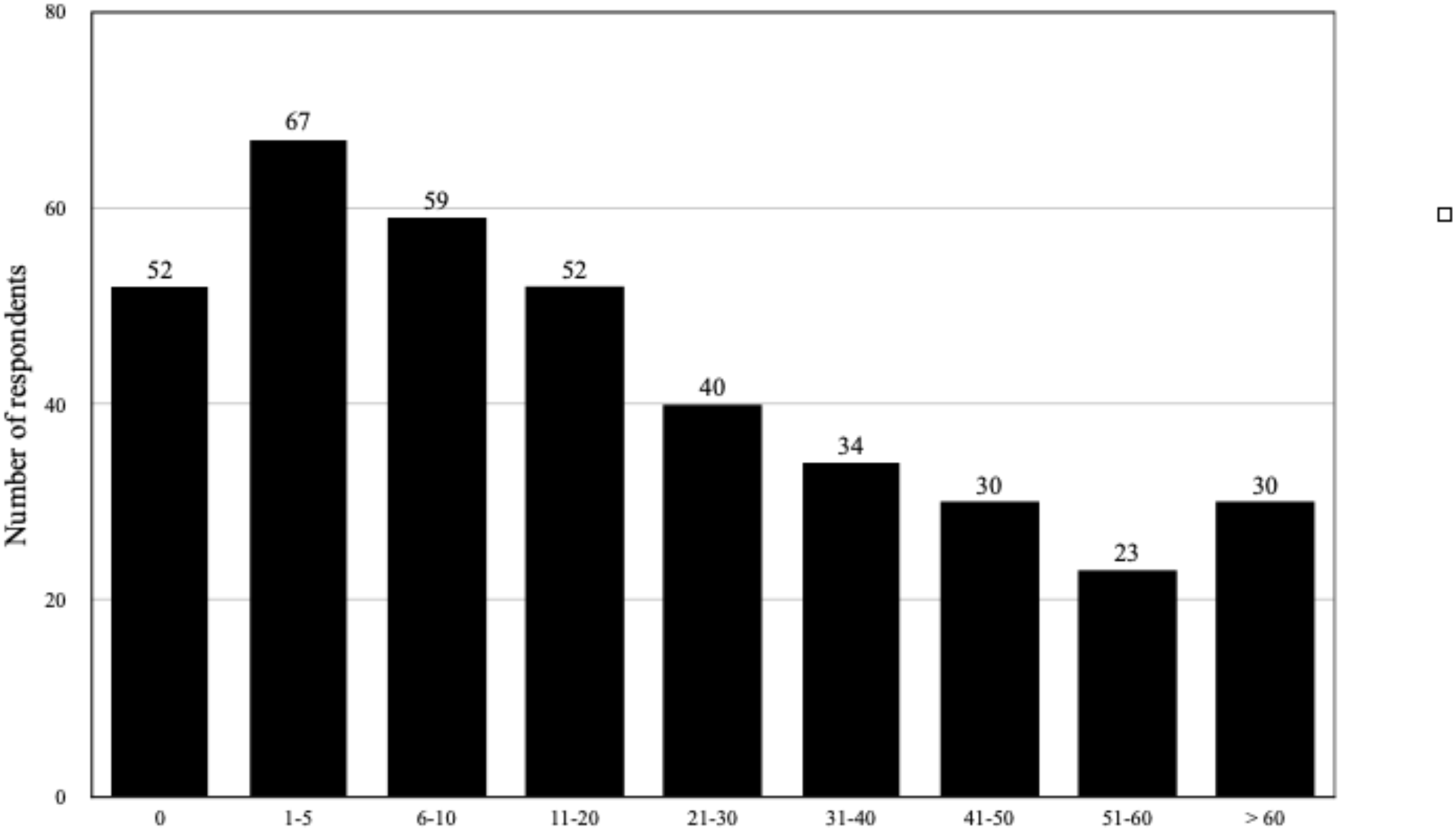
Impact on social network’s Score.

**Table 6:** Impact on Social Networks Composite of Severity by Incidence.

| Impact on Social Networks |  |  |
| --- | --- | --- |
| 1. How many people have you known who have been sick or died from HIV disease?<br>a. Family members or close friends who were/are **sick** from HIV<br>b. Family members and/or close friends who **died** from HIV<br>c. People you were less close to who were/are **sick** from HIV<br>d. People you were less close to who **died** from HIV | 1. Response-options for Family -members or close friends: 0, 1, 2, 3, 4, 5, 6-10, more than 10<br>2. Response-options for those “less close”: 0, 1, 2-5, 6-10, 11-20, 21-50, more than 50<br>3. The responses as to number of people were weighted by severity (Less-close-sick < Less-close-died < Close-sick < Close-died) and summed<br>4. The summed responses were collapsed into 9 groups | Mean=3.2<br>SD=2.5<br>n=387<br>Range=0-8 |
| Scores | n=387 | %age |
| 0 | 52 | 13.4 |
| 1-5 | 67 | 17.3 |
| 6-10 | 59 | 15.3 |
| 11-20 | 52 | 13.4 |
| 21-30 | 40 | 10.3 |
| 31-40 | 34 | 8.8 |
| 41-50 | 30 | 7.8 |
| 51-60 | 23 | 5.9 |
| >60 | 30 | 7.8 |

These results show that the experience of HIV-illness personally and socially varied markedly, with likely significantly different effects on the individuals affected. In the distribution of impact on social networks using a composite severity-by-incidence variable, a minority had known no-one in their social networks to have been sick or to have died from HIV-illness, and, numbers known related negatively to increasing severity-by-incidence; however, there was a spike at the end of the distribution in those with high severity-by-incidence, indicating a minority with severe experiences of personal illness and/or social loss from HIV who may form a specific subset with particular features and associated medical needs, and are distinctly different from those at the other end of the spectrum having had no such losses.

### Age-related differences in length and impact of HIV-infection

The relationship between age and length of HIV-infection was not uniform: the 40-49 age-group reflected a saddle-point, showing a significant increase in length of infection, with those younger than 40 more likely to have had HIV for less than 5 years, suggesting a wave of new infections in this group from 2004 onwards. Thus, for analyses, we compared the groups aged 20-39 with those 40-59 and, for length of infection, those having HIV for 5 years or less versus those for over 5 years. Using a composite variable of severity and incidence, the older group reported significantly more experience with mental and/or physical problems due to HIV or its treatment (Kendall’s tau-c=0.15; approx. T=3.08; p=0.002), as well as with other chronic conditions unrelated to HIV (tau-c=0.13; approx. T=3.03; p=0.002). Where reported, all probability-values are two-sided unless specified otherwise.

Overall, there was also a very strong, positive relationship between age and impact of HIV on social networks (χ^2^_df=8_=55.66, p<0.001), with some differences by severity-incidence across the proximity-hierarchy (family/close and died: χ^2^_df=7_=62.8, p<0.001; family/close and sick: χ^2^_df=7_=25.7, p=0.001; less close and died: χ^2^_df=6_=56.8, p<0.001; less close and sick: χ^2^_df=6_=19.7, p=0.003); this generational difference was particularly evident in knowing people who had been close/family and died (Kendall’s tau-c=3.01; approx. T=7.97; p<0.001). A similar, but stronger, association was seen with greater length of infection and impact of HIV-infection on social networks (χ^2^_df=8_=149.02, p<0.001). In light of the results below, these reflect significant differences in experience between generations as defined here that affect the primary research-questions.

### Access to physician-services

Although a condition of eligibility to respond to the survey was that the respondent had access to physician-services for HIV-treatment, if respondents stated that they did not have such access, they were asked to go to the end of the survey to answer the questions on ethnicity, residence, and provenance. Analysing those with reported access compared to those with reported lack of access or no-response (n=39, 10.9%), Fisher’s exact tests showed significant differences in access to services by age (χ^2^_df=1_=15.82, p<0.01) in that 29% (n=16/55) of participants younger than 30 years of age were not accessing services, which was three times the proportion of those 30 years or older (10%, n=37/368). Twice the proportion of women (24%, n=11/46) than men (11%, n=42/377) were not accessing services (χ^2^_df=1_=6.1, p<0.05), and more than twice the proportion of heterosexual men (24%, n=8) were not accessing services compared to gay men (10%, n=34; χ^2^_df=1_=6.27, p<0.05). No notable differences in access to physician-services were identifiable by language, length of time with HIV, where participants were currently residing, migration-status, or funding-type, specifically between “self-funded”, “employer”, “government”, and “combined self-employer”.

### Patients’ perception of, and response to, the quality of physician-service

To the authors’ knowledge, there are no psychometrics of the characteristics of patient-commitment to treatment and the issues that may affect such commitment. As an initial step towards such, we focused on respondent-physician communication (regardless of whether the HIV-services the respondents used were integrated — i.e., nurse-led or including psychologists), respondent-satisfaction and -perceptions of, e.g., physician-competence, and on behavioural outcomes such as change of physician due to disagreement. Fourteen questions (Table 7) covered respondent-satisfaction and relatedly significant domains, specifically: physicians’ perceived (1) communication-attributes; (2) relational conduct; (3) technical skill/knowledge; (4) the physician’s personal qualities; and (5) availability/accessibility.

**Table 7:**
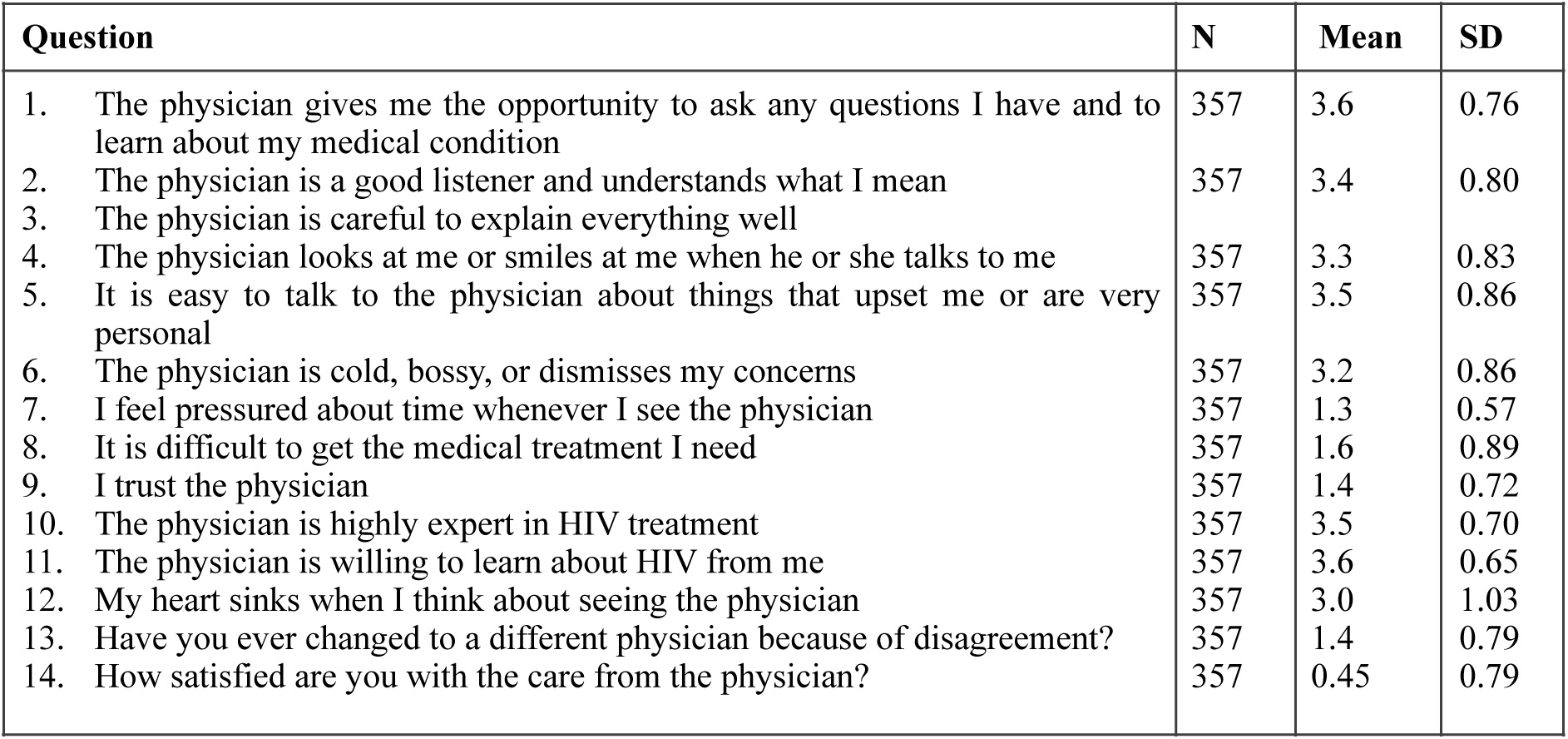
Questions regarding perception of, and response to, the quality of physician-service.

These questions were considered theoretically to reflect characteristics of the patient-physician interactions that affect patients’ commitment to HIV-services and eight of these questions dealt with respondent-perceptions of both positive and negative aspects of their physicians and their services provided. Referring to Table 7, there were four response-options for Questions 1-12 (e.g., from “always” to “rarely or never”; “very easy” to “very difficult”; “very willing” to “not at all willing”; etc.), 7 response-options for Q13 (from “never” to “6 times or more”), and 5 response-options for Q14 (from “completely satisfied” to “very dissatisfied”); all response-options were considered to be spline ordinal. The response-distributions of these variables were highly skewed; the least so—thus providing the greatest ability to differentiate across the spline-ordinal categories—were Q5 (indicating ‘clinical intimacy’; skewness=-0.91, SE=0.13) and Q11 (skewness=-0.57, SE=0.13). Due to significant skew, for analysis of reported satisfaction (Q14), we amalgamated the lowest three of 5 response-options (comprising the lowest 14.5% of the distribution of satisfaction) into “low satisfaction”; the remaining upper two response-categories reflected “moderate” (33.6%) and “high” satisfaction (the highest 51.9% of the distribution).

For respondents accessing physician-services for HIV, more rated their physicians positively than negatively. 84.3% (n = 301) reported that the physician was “often” or “always” a good listener, compared to 15.7% (n=56) reporting “rarely/never” or “sometimes”. 85.7% (n=306) reported that the physician makes eye contact and smiles “often/always”, and 14.2% (n=51) “rarely/never” or “sometimes”. 81.8% (n=292) reported that it was “quite/very easy” to talk to the physician about things that were personal or upsetting (“clinical intimacy”), and 18.2% (n=65) that it was “quite/very difficult”. 81.2% (n=290) reported that the physician was “rarely/never bossy”, and 18.8% (n=67) reported the physician was “sometimes/always” bossy.

However, there were noticeable age-differences as well: respondents aged 20-39 years rated their physician significantly lower in communication-skills than those aged 40-59 years (*F* (1, 354)*=*4.811, p=0.05) and were more likely to experience their physicians as cold, bossy, or dismissive (χ^2^_df=3_=14.346, p=0.002); similar findings were seen with variables of perceived time-pressure, and the experience of heart-sink. Significantly more of the older group also reported high clinical intimacy than the younger group (75%, n=104 vs 86%, n=188; χ^2^_df=1_=6.25, p=0.012) yet slightly more of the older group also reported having changed physician due to disagreement (Kendall’s tau-c=0.102, approx. T=2.2, p=0.028); this may have been due in part to the effect of an outlier group who had changed physician more than once and were older, possibly related to greater experience of HIV-impact or -illness, but it may also have been due to the younger group’s lesser experience of physicians and their lower likelihood of ever having taken HIV-medications (χ^2^_df=1_=16.72, p<0.001) and, for those who had, of having taken it significantly shorter time (χ^2^_df=4_=66.67, p<0.001) and having experienced fewer medication-toxicities (Kendall’s tau-c=0.511, approx. T=9.914, <0.001).

The findings above were paralleled by those with length of HIV-infection (e.g., the longer-infected were more likely to have changed physician due to a disagreement: Kendall’s tau-c=0.259, approx. T=5.442, p<0.001) and a comparison of the statistic-sizes indicated that the generational effect was most likely to be due to a greater average length of infection in the older group, indicating the effects of greater experience over time of HIV-illness on the longer-infected group (χ^2^_df=4_=14.6, p=0.006) as well as the impact of HIV on social networks (χ^2^_df=8_=149.02, p<0.001).

The reporting of any experiences of mental or physical illness due to HIV and/or its treatment varied widely and related positively to both increasing age (directional Somers’ d=0.11, approx. T=2.68, p=0.007) and length of time with HIV-infection (directional Somers’ d=0.18, approx. T=4.83; p<0.001).

From this set of questions, we extracted factors using Principal Components Analysis with split-sample validation (Table 8). Of these factors, only the Physician-Disrespect/Inaccessibility/Time-Pressure factor had a less-than-good performance, indicating an impure latent variable, as its variance explained initially was reduced by 6% to 57% following the removal of 8 outliers during split-sample validation; its constituent variables correlated only moderately: ‘cold-bossy-dismissive’ with ‘time-pressure during appointments’ (⍴=0.36) and ‘difficulty getting needed treatment’ (⍴=0.45); the latter two variables correlated similarly (⍴=0.44).

**Table 8:** Factors extracted through Principal Components Analysis.

| Variable | Items included | Factor-characteristics |
| --- | --- | --- |
| Length-Medication | 1. How long have you known that you have HIV?<br>2. How long in total have you taken medication for HIV? | Eigenvalue=1.7<br>Variance explained: 87%<br>Communalities=0.87<br>Cronbach’s $\alpha=0.9$<br>Component-loadings: 0.93 |
| Length-Impact | 1. How long have you known that you have HIV?<br>2. Composite: Impact on Personal Networks (see Table 4) | Eigenvalue 1.6<br>Variance explained 81%<br>Communalities >0.8<br>Cronbach’s $\alpha=0.78$<br>Component-loadings: 0.90 |
| Patient-Passivity | 1. Decisions about my medical care should be left to a physician<br>2. The people who benefit most from medical care are those who do what the physician tells them to do | Eigenvalue=1.4<br>Variance explained: 71%<br>Communalities=0.71<br>Cronbach’s $\alpha=0.60$<br>Component-loadings: 0.84 |
| Pragmatic Optimism | 1. I have a positive attitude towards my health<br>2. How good my health is in the future depends on what I do about it now | Eigenvalue=1.4<br>Variance explained: 72%<br>Communalities=0.72<br>Cronbach's $\alpha$=0.63<br>Component-loadings: 0.85 |
| Perceived Service-Quality | 1. How satisfied are you with the care from the physician?<br>2. The physician gives me the opportunity to ask any questions I have and to learn about my medical condition<br>3. The physician is a good listener and understands what I mean<br>4. The physician is careful to explain everything well<br>5. The physician looks at me or smiles at me when he or she talks to me<br>6. It is easy to talk to the physician about things that upset me or are very personal<br>7. The physician is cold, bossy, or dismisses my concerns<br>8. I trust the physician | Eigenvalue=5.1<br>Variance explained: 63%<br>Communalities >0.55<br>Cronbach's $\alpha$=0.92<br>Component-loadings: 0.75-0.86 |
| Physician-Engagement | 1. The physician allows me to ask any questions I have and to learn about my medical condition<br>2. The physician is a good listener and understands what I mean<br>3. The physician looks at me or smiles at me when he or she talks to me<br>4. The physician is willing to learn about HIV from me | Eigenvalue=2.6<br>Variance explained: 66%<br>Communalities >0.54<br>Cronbach's $\alpha$=0.83<br>Component-loadings: 0.73-0.84 |
| Physician-Credibility | 1. The physician is careful to explain everything well<br>2. I trust the physician<br>3. The physician is highly expert in HIV treatment | Eigenvalue 2.05<br>Variance explained 68%<br>Communalities >0.62<br>Cronbach's $\alpha$=0.78<br>Component-loadings: 0.79-0.87 |
| Physician-Disrespect/<br>Inaccessibility/<br>TimePressure | 1. The physician is cold, bossy, or dismisses my concerns<br>2. It is difficult to get the medical treatment I need<br>3. I feel pressured about time whenever I see the physician | Eigenvalue 1.7<br>Variance explained 57%<br>Communalities >0.52<br>Cronbach's $\alpha$=0.59<br>Component-loadings: 0.72-0.79 |

### Satisfaction, trust, and their predictors

Respondents’ trust in their physician was high on average: 357 (85%) of the sample answered this question, of which 59.7% (n=213) reported that they could trust the physician “completely”, 31.3% (n=111) trusted the physician “mostly”, and 9.2% (n=33) “somewhat/not at all”. Satisfaction was slightly lower: 366 (87%) of the sample responded to this question, of whom only 190 (44.9%) were completely satisfied with the ‘care’ from their HIV-physician; 129 (29.1%) were quite satisfied, 42 (9.9%) somewhat dissatisfied, 7 (1.7%) quite dissatisfied, and 4 (0.9%) very dissatisfied; thus, 12.5% or 1 in 8 had some degree of dissatisfaction. There were no differences identifiable in satisfaction by age, sexual orientation, language spoken, ethnic group, or migration-status, nor indeed by length of HIV-infection.

Given its salience in the literature, predictive models of satisfaction were developed first through decision-tree analysis (DTA) as logistic regression was not an optimal choice due to the non-normality of residuals. We determined which of the variables comprising the Perceived-Service-Quality factor specifically were best able to predict reported satisfaction itself but we also included the five other questions in the question-set due to their communalities being less than 0.5; only individual questions were used as inputs. Further, when the primary predictor was a factor or composite, we excluded from confirmatory analyses all variables that were constituents of the primary predictive factor or composite. Models that excluded all variables contributing to the Perceived-Service-Quality factor showed only a poor risk-estimate (0.43) and were not pursued.

In repeated overall models (risk-estimates: 0.26-0.29 with re-substitution and 0.29-0.32 with cross-validation), the best predictor of respondents’ “satisfaction” with physician-services for HIV was Perceived Physician-Credibility (χ^2^_df=4_=243.92; p<0.001), a factor consisting of the perceived quality of physician’s explanations, trustworthiness, and expertise in HIV-treatment. In low physician-credibility (percentiles 9.6 - 23.1), lowest satisfaction was predicted primarily by those in the upper 30% of the Physician-Disrespect/ Inaccessibility/Time-Pressure distribution, a factor consisting of physician-disrespect, treatment-inaccessibility, and/or perceived time-pressure during appointments. In moderately low perceived Physician-Credibility (percentiles 23.2 - 38.1), higher satisfaction was predicted by “always” being given the opportunity to ask questions. In moderate physician-credibility (percentiles 38.2 - 62.7), higher satisfaction was predicted by the respondent’s positive attitude to his/her health and, in those with less than a permanently positive attitude, any problems with Physician-Disrespect/Inaccessibility/Time-Pressure. In highest physician-credibility (percentiles 62.8 - 100.00), satisfaction was affected negatively by perceived time-pressure during appointments or, in the absence of perceived time-pressure, by ever having had the experience of very serious HIV-illness. In this overall model (Figure 3), the best predictors of *high* satisfaction was a respondent being in the highest 37% of the distribution of reported Physician-Credibility (87.1% correctly predicted), increasing if having “rarely” or “never” experienced time-pressure whenever the respondent saw the physician (90.7% correctly predicted), and additionally never having had “very serious” problems due to HIV or its treatment (96.3% correctly predicted). The best predictor of *low* satisfaction (“somewhat” to “very” dissatisfied: 79.4% correctly predicted) was a respondent’s being in the lowest 9.6% of the distribution of perceived Physician-Credibility.

**Figure 3:**
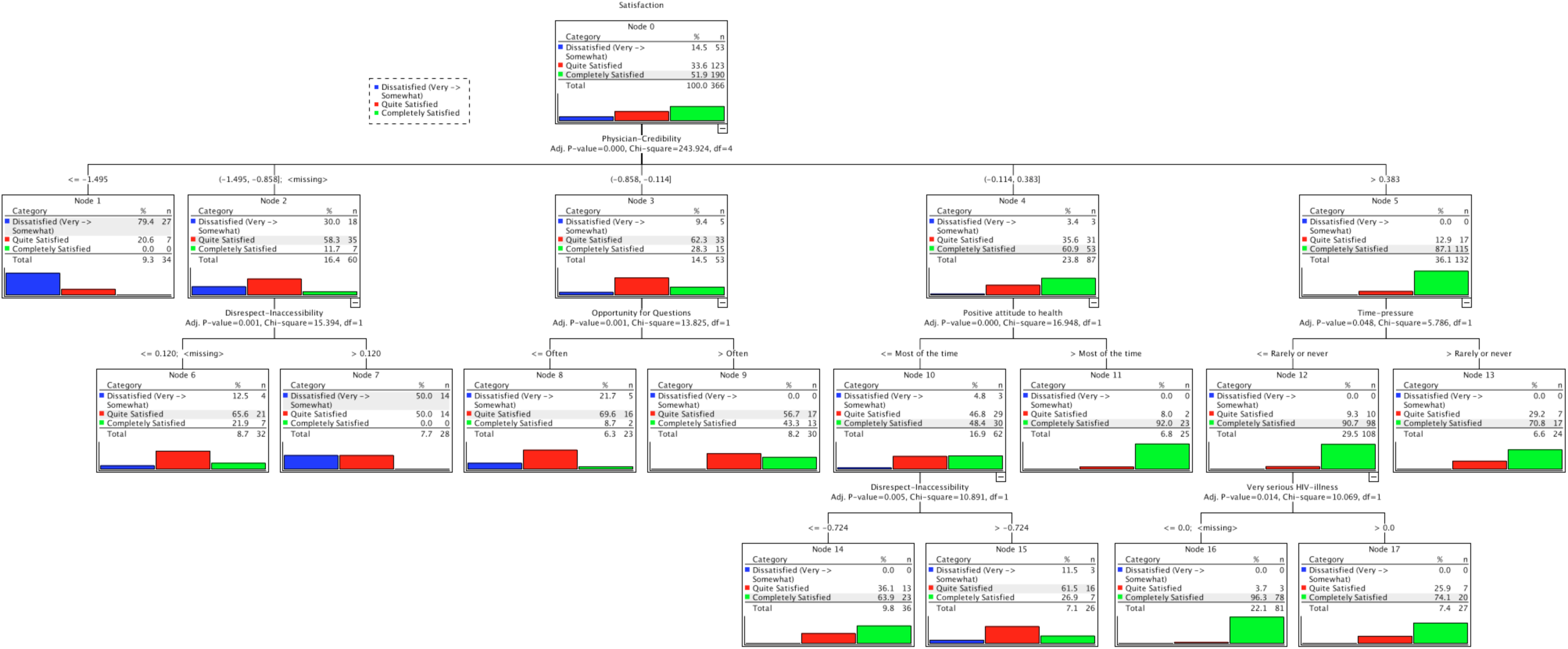
Overall DTA-model predicting Satisfaction.

Creating models to predict specific regions of the satisfaction-variable’s distribution only, the model providing the best predictor of *high* satisfaction (92% correctly predicted) was “complete” trust in the physician (comprising the upper 59.7% of the distribution of that variable), who “always” provides the opportunity both to ask any questions the respondent has and to learn about his/her medical condition (comprising the upper 69.2% of the distribution of that variable), who appears to be “highly” expert in HIV treatment (comprising the upper 68.6% of the distribution of that variable), and with whom the respondent finds it “very” easy to talk about “things that upset me or are very personal” (comprising the upper 45.7% of the distribution of that variable). In contrast, the model providing the best predictor of *low* satisfaction (88% correctly predicted) identified those who trust their physician “somewhat” or “not at all” (comprising the lowest 9.2% of the distribution of trust). Jointly, these models all performed well (risk-estimates: re-substitution=0.26-0.27, cross-validation=0.28-0.34) and provided convergent evidence as to the identification of the primary predictors of overall, high, and low satisfaction, which was primarily respondents’ trust and which may be the primary constituent of the Physician-Credibility factor.

Given the role of respondents’ trust in determining satisfaction, an independent DTA to find the predictors of trust indicated that a single model resulted, with good performance-characteristics (risk-estimates: re-substitution=0.261; cross-validation=0.3). Briefly, this model showed that the predictor of highest trust was the level of Physician-Engagement factor, high Engagement predicted by having high Pragmatic Optimism; the predictor of middling trust was, relatedly, a low endorsement of the statement “I believe that what HIV will do to me in the future is completely out of my control” (indicating low self-efficacy). The predictor of lowest trust was low Physician-Engagement in a physician who was perceived to be less than “highly expert” in HIV-treatment. Removing complex factors resulted in the primary predictor of trust being the physicians’ listening-skills. However, while the effects of poor listening-skills were seen to be mitigated slightly by physicians’ perceived expertise, it was not enough to increase the respondents’ trust, showing that listening-skills are more important to the patient than perceived expertise.

Thus, satisfaction is predicted primarily by Physician-Credibility, which is affected by variables such as physicians’ attitudes, time-pressure, access to treatment, and communication-skills, as well as respondents’ attitudes to health and their experience of HIV-related illness. Focusing on specific areas of satisfaction showed that respondents’ trust was key.

### Heart-sink and its predictors

Most of the 357 (84.4%) respondents to this question “rarely or never” felt heart-sink when they thought about seeing their physician, but this left a significant minority who “sometimes” (17.9%), “often” (7%), or “always” (3.9%) felt it. Heart-sink correlated with satisfaction significantly (⍴=-0.45; p<0.001) and there were also age-related associations with the experience of heart-sink: it was higher in participants aged 20-39 years, with 36% of participants experiencing heart-sink always/often compared to those aged 40-59 years with 25% of participants (χ^2^_df=1_=4.86; p=0.028). Heart-sink was not related to gender, length of time with HIV, migration-status, language spoken (between English and Spanish), ethnic-group identification (between Britain and the rest of Europe, nor between American North and American European), nor sexual orientation in males—there were too few female participants who identified as lesbian or bisexual to compare sexual orientation in females.

The most potent predictors in initial models of heart-sink were complex factors consisting of relational and related contextual variables, such as clinical intimacy, trust, warmth, and time-pressure in physician-appointments. Removing these complex factors for the purposes of clarity, the best model’s predictor (risk-estimates: re-substitution = 0.19, cross-validation = 0.28) was respondents’ trust in the physician (χ^2^_df=2_=71.24; p<0.001). Predicting those whose hearts sank “often” or “always” was determined best (82% correctly predicted) by those who trusted their physician “somewhat” or “not at all”, and (72% correctly predicted) by those who “somewhat” or “mostly” trusted their physician and who were in the *lower* 58% of the distribution of the Length-Impact factor.

The best predictors of having one’s heart sink “rarely” or “never” were: (1) reporting very low or no anxiety when reading or hearing about HIV (“rarely or never”; 96% correctly predicted); and (2) in those with at most low anxiety (“sometimes, rarely or never”), who also lay in the 38.7% of the distribution between the 33.1st and 71.8th percentiles of Pragmatic-Optimism factor (96% correctly predicted). When all physician-respondent relational variables were excluded (Figure 4), the strongest predictor of heart sinking “rarely or never” (89% correctly predicted) in the best model (χ^2^_df=3_=56.9; p<0.001; risk-estimates: re-substitution = 0.25, cross-validation = 0.28) was in those who “rarely or never” felt anxious or depressed when hearing or reading about HIV; likewise, the best predictor of one’s heart-sinking “always/often” was feeling anxious or depressed when hearing or reading about HIV (76% correctly predicted). Thus, the experience of a heart-sink physician was driven by anxiety and mediated by lesser trust in that physician.

**Figure 4:**
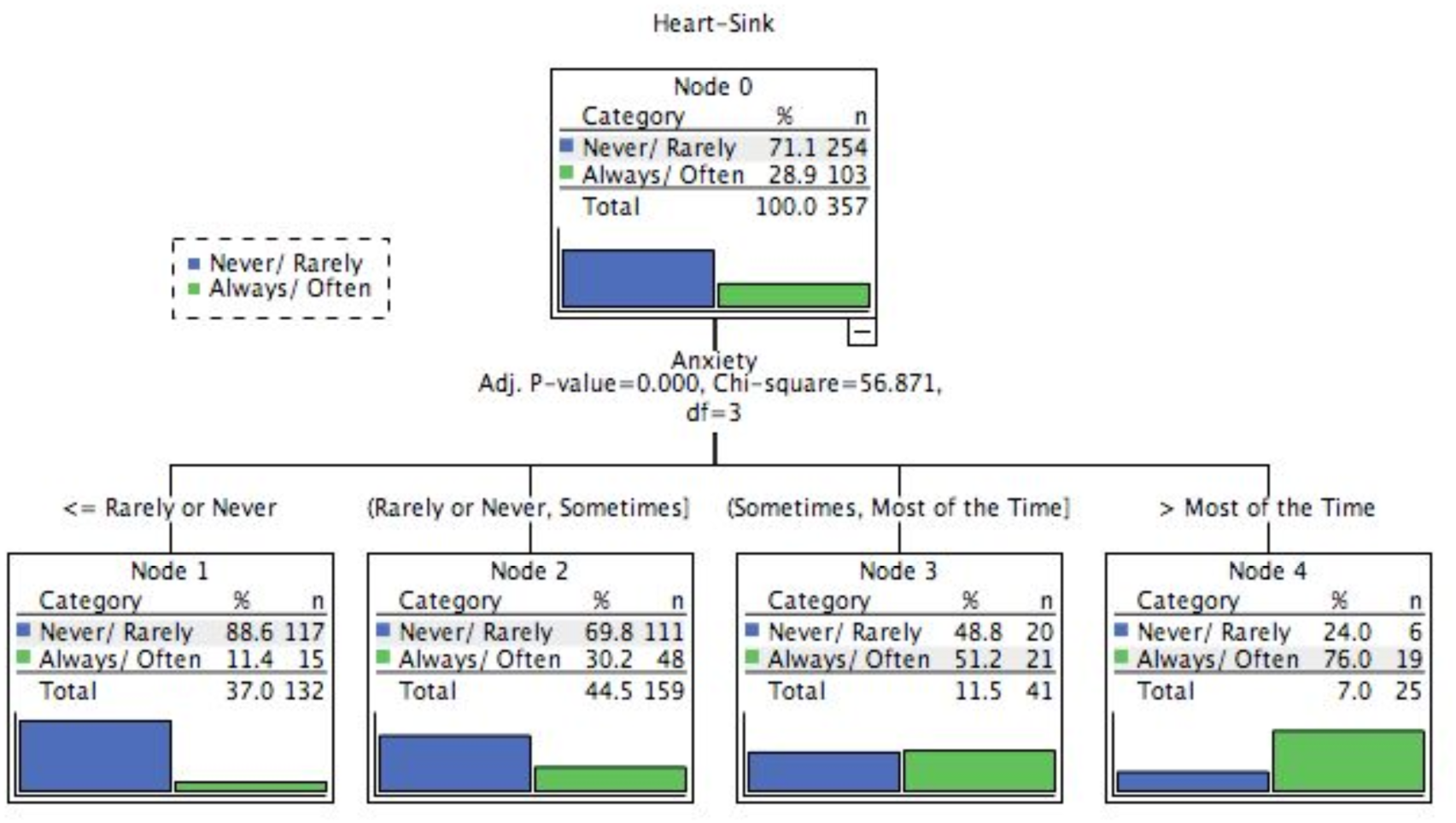
DTA-model predicting Heart-Sink.

Anxiety’s strongest positive correlates were the “Patient-Passivity” factor (i.e., ‘I prefer clinical decisions to be left to my physician’) and a sense of helplessness/fatalism, as indicated by agreeing with the statement “I believe that what HIV will do to me in the future is completely out of my control” (both, ⍴=0.4; p<0.001). Similarly, respondents’ obedience and passivity correlated negatively with satisfaction (⍴=-0.31), Perceived Service-Quality (⍴=-0.32), Physician-Credibility (⍴=-0.34), and positively with heart-sink (⍴=0.34). Thus, respondents lacking a sense of control over their condition and its treatment were less likely to trust, to view positively, and to be satisfied with their physician-services for HIV. This has significance for the model of the ‘good’ patient traditionally preferred by physicians.

### Disagreement sufficient to cause a change of physician

Most respondents answered this question (84%), of whom 69.2% had never had a disagreement with their physician sufficient to cause them to change their physician; 20.7% had done so once, 7.3% twice; 1.7% three times, and 1.1% four times (Table 3). Thus, almost a third of the sample (30.8%) had had a disagreement with their HIV-physician such that it caused them to change physicians at least once. Such disagreement was not related to gender, migration status, language spoken (between English and Spanish), ethnic group identified with (between Britain and the rest of Europe, nor between American North and American European), sexual orientation in males, nor satisfaction. This question was consistently independent in Principal Component Analyses from other items. Its highest correlates related to the length of time since infection, the impact on the respondent’s social networks, and whether the respondent had ever experienced medication-toxicity.

The proportion of respondents reporting disagreement with the physician sufficient to change physician was higher in participants aged 40-59 years, with 13% of participants experiencing such disagreement twice or more, compared to those aged 20-39 years with only 5% (χ^2^_df=2_ = 6.59; p = 0.037). Disagreement was also higher in participants infected with HIV for 11 years or more, with 35% experiencing disagreement once, and 22% twice or more, compared to those with HIV for 3 years or less, with 16% and 6% respectively (χ^2^_df=2_ = 37.08; p < 0.01).

The strongest predictor in the best predictive model of disagreement likely to cause a change in physician (risk-estimates: re-substitution: 0.29; cross-validation 0.33) was “Length-Impact” (χ^2^_df=2_=45.6; p<0.001) of HIV-infection. Convergent analyses showed that the primary contributor to this effect was not the length of time infected with HIV but the impact of HIV on social networks, relating primarily to incidence (as in number of people), not severity (as in closeness and death, rather than illness). The best predictor of any such disagreement (74% correctly predicted of any disagreement, and also 48% correctly predicted having two or more such disagreements) was being in the highest 19% of the Length-Impact factor’s distribution and having had more than two instances of changes in medication due to medication-toxicity (the latter also being a primary correlate: ⍴=0.29, p<0.01 just less than length of HIV-infection ⍴=0.294, p<0.01). The second-strongest predictor of any such disagreement (68% correctly predicted) was being in the 29% next highest in the Length-Impact factor’s distribution while also finding it “quite” or “very” difficult to talk with the physician about things that are personal or upset the respondent (i.e., poor/low clinical intimacy). Thus, the contribution to disagreement causing change in physician was the number of people in the respondent’s network who had suffered from HIV, the length of the respondent’s own infection, and the experience of changing medication due to toxicity; together, these would point to a convergence of emotional impacts on the respondent. It is notable that the experience of HIV-illness itself was not related to such disagreement.

Similarly, the best predicted in terms of a *lack* of any such disagreement was being among those in the lowest 50% of the distribution of Length-Impact while also never having taken HIV-medication (93% correctly predicted); the second-best predicted were in the same region of the distribution on the Length-Impact factor, had taken HIV-medication, but being in the lower 50% of the distribution of the Physician-Disrespect/ Inaccessibility/Time-Pressure factor (90% correctly predicted). It is noteworthy that, even in those with the lowest Length-Impact group who also took medication but experienced Physician-Disrespect/Inaccessibility/ Time-Pressure, 40% of people who changed physician because of disagreement were correctly predicted. Further clarifying this picture was the third-best predictor of never experiencing disagreement sufficient to cause a change in physician, which was being in the middle group (between 53rd and 81st percentiles) of the Length-Impact Factor, who found it “quite” or “very easy” to talk with the physician about things that were personal or upset the respondent, and who have never had any mental or physical problems due to HIV (83% correctly predicted). This distinctive variable of changing physician due to disagreement may be a proxy for problems in physicians’ ability to relate to patients’ emotional consequences of HIV, such as anxiety and trauma. Thus, these indicate that likely primary causes of changing physicians following disagreement are due to the effects of trauma from HIV-losses and other causes^7^ in tandem with the physician’s poor relational skills; this has significance for losses to follow-up and patients’ disengagement from physician-services. This also supports existing calls for trauma-informed services^8^.

## Discussion

In order to inform the future design and delivery of health-services for people with HIV, as well as other chronic conditions, this study explored the characteristics of HIV-infected people’s relations to physician-services for HIV-infection and possible reasons as to why commitment to treatment and retention of physician-services may diminish or disappear. This was done by assessing perceived characteristics of both respondent and physician, relational issues, and contextual issues (such as impact on social networks, illness-experience, and perceived time-pressure in appointments). The development of the study was informed by grounded theory and participative research, based on the primary investigator’s and other patients’ direct experiences and in line with the theoretical literature, which enabled the inclusion of novel questions (e.g., bossiness; heart-sink; conflictual disengagement) and issues that are typically excluded from research but feature significantly in patients’ experiences. The survey’s degree of success was facilitated by the interface-design, despite significant complexity and response-conditionalities, and particularly by its ability to rely for its delivery on an online, community-focused platform (www.thebody.com) with its access to diverse parts of the community that would otherwise be unavailable. While this means had instructive problems associated with it (e.g., affecting language-capture in this instance), such an asset to community-focused research is invaluable and irreplaceable. As a result, a random-consistent sample was achieved in a modest sample-size that provided data of significant utility. Further, although individual statistical results are reported here for the sake of completeness, and probability-values are Bonferroni-adjusted where indicated, the focus is not, and should not be, on specific results, as this is an exploratory study, but is instead more on convergent analysis in order to identify potentially robust findings and their inter-connections in the construct-space for the purpose of future, (dis-)probative research.

This sample showed a remarkable degree of loss in social networks due to HIV-disease and the most significant HIV-associated illnesses reported as mental ones, particularly depression and PTSD; other studies have found that almost half of HIV-positive people suffer from mental disorders^9^, notably depression and (especially, complex) PTSD or related disorders of extreme stress^10,11^. The prevalence of such disorders is of little surprise given the relationship of the risk of HIV-acquisition to social marginalisation and related poverty and substance-abuse, the prevalence of stigmatisation and resulting discrimination, illness itself from HIV and/ or its treatment, the perceived fatality of the disease prior to availability of effective antiretrovirals, and—most notably—the impact on social networks. These are not single-event stressors but traumas that have saturated patients’ lives over time. Mental illnesses, including problematic drug-use-behaviours and dependency-conditions, remain salient behavioural drivers of the acquisition of HIV^12^ but also affect the ability to manage and recover long-term from HIV-illness given the availability of ART^13^.

These findings speak to the effects of mental, social, and behavioural issues on the way in which people with HIV view health-services, affecting in turn their linkage to health-services^14^, the initiation and continuity of necessary treatment^15–17^, treatment-cascades^18,19^, primary and secondary prevention^20–22^, the speed of disease-progression, and survival^23–25^, and thus the effectiveness and efficiency of services in helping people to get well and stay well^26^. In the general population, the effect of mental disorders is greater than the effects of major physical disorders, and they interact when comorbid^27^; this is likely to be even more so in HIV, in which mental illness is known to reduce, significantly and directly, the role-related functions and capabilities of people with HIV^28^. Further, as the pandemic persists, the ageing population of people with HIV is proving to be in particularly greater need of health-services, as the effects of comorbid cardiovascular, renal, osteoporotic, pulmonary, cognitive and other neural disorders are likely to interact with other physical, mental, and social causes of age-related vulnerability—including frailty and loneliness^29^—and therefore self-sufficiency among the aged.

While an idea of the needs and perspectives of patients as whole people now imbues health-services’ public relations, it has yet to affect practice significantly^30^, not least in the fact that physician-centred services typically continue to deprecate mental and social problems. The physician-centred attitudes that underlie such services also affect service-formulation, maintaining models of health-services that are outdated^31^. The deprecation of mental and social aspects of illness results in the use of service-providers of inadequate competence, such as General Practitioners (physicians) with no training in psychopathology or social work^32^, low-level counsellors with minimal training in a restricted skill-set, and even unsupervised non-professionals with lived experience leading drug-treatment services in HIV-clinics. Most (or even all) HIV-clinics internationally deliver balkanised, physician-centred disease-services rather than integrated, comprehensive, and patient-centred health-services, and thus fail to be optimally health-effective^33,34^. Such balkanisation was not always the case in HIV-services: integrated and co-productive services existed prior to the availability of effective ART in 1996, after which time the standard, physician-centred model of disease-services resumed control and service-quality and -effectiveness suffered^35^, at least in the global West.

The idea of the needs and perspective of patients as whole people has yet to affect not only practice but also how scientific and technical research is both conceptualised and reported: for example, the research-literature reports on physicians’ “retention of patients”^36,37^ where patients are characterised as objects retained as an action-step in the HIV-treatment cascade^38^. In contrast, a frame in this paper is of patients’ “retention of services”, with patients characterised as self-determining individuals with a complex personal life that affects their ability and willingness to commit to and engage with health-services, thereby maximising services’ health-effectiveness. Only the latter construction provides potential for optimising services by meeting patients’ needs as whole, engaged people, rather than as infected bodies.

This study addresses the concept of ‘satisfaction’ in terms of its nature, predictors, and adequacy. Currently, service-quality and patients’ likely future engagement with services are considered to be measured in part by patients’ satisfaction with health-services generally and with their treating-physician specifically^39^. This study finds that respondents’ satisfaction is determined mostly by physician-characteristics, such as perceived physician-credibility, communication-skills, and technical expertise, but also by respondents’ trust and sense of pragmatic optimism. Perceived physician-credibility and particularly trust in physicians was highly important and a non-linear effect might be expected, in that those with less negative experience of HIV on their health and social networks would have less reason for any perceived credibility to have been tested. It is also reasonable to suggest that patient-trust is likely to be different for acute vs. chronic and/or complex conditions. How well the physician then responds to a patient’s more severe and complex problems would be key; paternalistic interventions like physicians’ solely providing reassurance, a tool to reduce their own anxiety but also reducing patients’ readiness to report physical or mental complaints, can be counterproductive for patients when underlying issues are not identified and addressed.

That said, there are scientific problems with the construct of ‘satisfaction’, not least that there is no scientific evidence to indicate a necessary or sufficient determinative relationship between patient-satisfaction and health-outcomes (i.e., ‘quality’ from the patients’ perspective). Logical analysis argues against such a relationship, particularly in risk-management based on weakest-link analysis of cases such as that of Harold Shipman^35^, a generalist physician whose then-current patients rated his services as so highly satisfactory that they attested to that during his trial — in which he was found guilty of murdering over 250 of his other patients. Thus, the lack of ecological validity of satisfaction as an index of the quality of physician-service is obvious if unnecessary mortality, let alone morbidity or malpractice, are outcomes to be avoided.

There are also wider methodological and conceptual problems with satisfaction and its measurement^40^, as with typical measures of “patient-experience”, as it represents a relational stance based on emotional ephemera rather than an applied and informed opinion based on objective evidence of health-effectiveness, and typically results in data having a skewed distribution with ceiling-effects and reduced discriminatory ability^41,42^. Despite satisfaction’s lack of validity, physicians have tried to measure it in HIV-services, resulting in instruments of additionally low psychometric and conceptual validity^43^. The confusion caused by consumerist and “managed-care” approaches to service-quality that lack a scientific understanding of concepts such as “satisfaction” has likewise given rise to the “Friends and Family Tests”^44,45^, which suffer from the same problems of validity.

Given the drivers of service-retention — this being fundamental to services’ health-effectiveness — it was important to identify the predictors of potential disengagement from services. In the present study, two pivotal variables other than satisfaction were explored that may assist in opening up future research on predictors of patient-commitment and services’ health-effectiveness: whether or not respondents’ hearts sink at the thought of seeing their physician and whether or not they had ever changed physician due to a disagreement. Modern understandings of the ‘good’ patient are of an ‘activated’ and engaged person with self-efficacy; the characteristics of empowerment and self-determination are not part of that picture and, while the idea of obedience and passivity as marks of a ‘good’ patient is increasingly obsolete in Western clinical literature^46^, in practice, patients are often termed “heart-sink”^47–49^, “difficult”^50–52^, or even “hateful” patients^53^ when they are non-acquiescent or even “ungrateful”; when they present complaints that are not purely physical or which are “excessively difficult” diagnostically; or when they seem to have emotional, hypochondriacal, somatoform, or “medically (i.e., *bio*medically) unexplained” characteristics — types of conditions falling within the rubric of psychiatry, which many physicians distinguish from ‘real’ medicine^56,57^[54.55] and, thus, not something that they should have to spend their valuable time on. This reflects a wider problem with physicians’ attitudes towards themselves and towards patients, both of which can affect patients’ commitment to service-retention.

Although we have been unable to find evidence that ‘heart-sink’ has ever been applied to physicians in the literature previously, patients also clearly experience heart-sink in relation to physicians. While the full characterisation of heart-sink physicians remains for future research, in this study, ‘heart-sink’ in respondents related primarily to anxiety and fatalism/helplessness when seeing their physician, and negatively to satisfaction. The fundamental predictors of heart-sink were distinct from those of satisfaction and reflected primarily patient-characteristics (anxiety, medical passivity, both length and impact of HIV-infection, and a lack of sense of control), rather than physician-characteristics. It would be reasonable to suggest that the use of the term heart-sink by physicians about patients is driven similarly by emotional problems such as anxiety and poor clinical self-efficacy within physicians.

The study noted that both satisfaction and heart-sink were independently predicted best by trust in the physician, while conflictual disagreement was not, although the latter’s predictors may suggest a loss of trust due to traumatic and/or chronic stress. The findings suggest that trust is earned by physicians’ engagement and relational skills, and is affected positively by respondents’ sense of pragmatic optimism. Trust is lost by the experience of time-pressure as well as by patients’ anxiety and fatalism, and both trust and satisfaction are lost with increased experience of HIV-illness and its negative effects on the respondent, such as medication-toxicity and traumatic loss through the impact of HIV on patients’ social networks. It is of particular interest to note that trust was negatively related to obedient passivity, suggesting a conditional acquiescence in ‘good patients’: while acquiescent patients may be physicians’ preferred type, acquiescence is thus distinct from trust.

The second driver of patients’ non-retention of services mentioned above, was the issue of changing physician due to a disagreement. The unique construct of ‘disagreement leading to a change in physician’ appeared to be conceptually orthogonal to satisfaction and may have relevance to the issue of patients’ commitment to treatment. Almost a third reported having changed physicians for this reason and conflictual disengagement from physician-services was predicted best by a higher impact of HIV on social networks, more negative experiences of mental and/or physical illness from HIV and/or its treatment—particularly changing medication due to toxicity (of which almost half the sample had had cause to switch medications for this reason)—along with poor ‘clinical intimacy’ and a greater sense of time-pressure, consistent with other research^56^. Conflictual disengagement was also more likely in older people and those who had had HIV longer, which may reflect generational differences in experience of HIV-illness and social loss, and thus respondents’ relations to health-services, but entails an increased risk among a group already at increased risk of age-related morbidities^57^. Such disengagement did not occur in relation to severity of HIV-related illness, the discrepancy pointing to the significance of whether or not physicians can actually address patients’ full health-needs or only the physical ones, and what happens when the social and mental aspects of illness are not competently addressed.

When patients’ trust in physicians’ ability to address adequately their illness-related needs is eroded, it may engender sufficient conflict—evident or not—for the patient to disengage from physician-services. While excellent relational, including communicational, skills in physicians, not least the ability to provide space for emotional intimacy and disclosure while protecting the patient from perceived time-pressure in the clinical encounter, are both essential and rare, they may be insufficient to compensate for an erosion of trust and credibility in the face of such social and mental consequences of HIV. These point clearly to means by which to increase trust in health-services, including a fully competent addressing of mental illness arising from complex personal histories, empowering patients by addressing passivity and fatalism, ensuring excellent communication-skills in clinicians, and ensuring maximally efficient and otherwise health-effective services.

The present research identifies three variables relevant to the question of service-retention — satisfaction, heart-sink, and conflict-driven change of physician; the latter two are actually more useful and informative than satisfaction, as both reflect issues within the patient rather than professionals’ solipsistic focus on service-variables that determine satisfaction. This makes more sense psychologically and provides greater latitude for intervention. In light of the problems discussed here, we propose that the cluster of concepts such as satisfaction, engagement, activation, etc., are inadequate to explain the acquisition and/or loss of engagement and retention of health-services for HIV. Rather than promoting consumerist indices such as ‘satisfaction’ while perpetuating physicians’ professional dominance and the health-ineffective, physician-centred model of services^31,58^, the discourse of research and practice should focus on concept-clusters reflecting patients’ mental and social burden(s), clinical intimacy, passivity-inefficacy, trust-distrust, and clinical conflict as determinants of patients’ engagement with, and commitment to, health-services.

To be health-effective, services have some absolute requirements, which are predicated upon patients’ access to, engagement of, and retention of health-services; mutually committed clinical relationships; and competent treatment of the full, biopsychosocial range of illness-phenomena^30,35^. Ethical concerns arise as to physicians’ lack of adequate competence to detect, evaluate, and address the range of mental and social issues associated with having HIV and the adequacy of services to address those in an integrated manner. This not only supports the role of nurse-practitioners as the primary clinician and portal to integrated services, but having clinicians such as psychologists address mental issues may actually protect physicians’ credibility and patients’ engagement and retention of services in the face of, e.g., difficult illness and adverse medication-experiences. However, fully integrated services would require a move from physician-centredness to patient-centredness in the design and delivery of services^31,35^, while the tension between the emotional needs of physicians for professional dominance^58^ on the one hand and the physical, mental, and social illness-needs of patients on the other indicates the scale of the obstacles to the re-development of integrated services. Thus, the health-effectiveness of services, and patients’ retention of them, may depend on which is considered to be more important: physicians’ needs or patients’ needs.

## Data Availability

All data produced in the present study are available upon valid request to the lead author.

## Acknowledgements

This study was made possible by support from: Thebody.com, New York (coding and hosting of survey); the University of Roehampton, London (ethics committee); the University of Warwick (access to statistical software); Podio.com, Gerrards Cross, UK (online platform for remote collaboration and data-storage); Prof. Jim Wiley, University of California San Francisco, USA (conceptual and statistical assistance); Cecilia Gomez, Ruben Dogliani, and Daniel Cambuzat (translation to and from Spanish). It is dedicated to the memory of Paul Blanchard (1964-2016), physician.

